# Adherence to the Eatwell Guide and associations with markers of physical function: A prospective analysis within the UK Biobank cohort

**DOI:** 10.64898/2026.04.27.26351814

**Authors:** Alex Griffiths, Sarah Gregory, Fiona C Malcomson, Kirstie Cronin, Jamie Matu, Louisa Ells, Oliver M Shannon

## Abstract

**Background:** The Eatwell Guide represents the UK’s principal healthy eating model and understanding whether adherence to UK dietary recommendations can attenuate age-related functional decline is essential to inform healthy ageing strategies.

**Methods:** In up to 157,457 participants from the UK Biobank, we explored cross-sectional and prospective associations between adherence to the Eatwell Guide and markers of physical function (grip strength, fat-free mass percentage, self-reported walking pace, and falls). Eatwell Guide adherence scores were derived from 24-hour dietary recall data (Oxford WebQ), and quantified using a graded, food-based scoring system. Differences between population subgroups including by age, sex, physical activity, and protein intake level were explored.

**Results:** Higher Eatwell Guide adherence was cross-sectionally associated with higher grip strength, greater fat-free mass percentage, higher odds of brisk walking pace, and lower odds of falls (all p<0.001). Prospectively, greater adherence was associated with attenuated fat-free mass decline (β=0.02, SE=0.001, p<0.001) and slower grip strength decline (β=0.01, SE=0.002, p<0.01). Higher adherence was also prospectively associated with greater odds of brisk walking pace (OR=1.02, 95% CI: 1.017-1.021, p<0.01), though this advantage attenuated over follow-up (EWG*Time: OR=0.998, 95% CI: 0.997-0.999, p=0.002). Higher adherence was prospectively associated with lower falls risk (OR=0.996, 95% CI: 0.995-0.998, p<0.001), with this protective association remaining stable over time (EWG*Time: p=0.89).

**Conclusions:** Higher Eatwell Guide adherence was associated with preserved muscle mass, modest attenuation of grip strength decline over time, and a reduced risk of falls, supporting its relevance for musculoskeletal health and physical function in ageing populations.

## Introduction

The number of adults aged 65 and over in the UK is projected to almost double in the next 50 years to > 22 million (Office for National Statistics, 2024). As the population ages, the prevalence of physical decline is expected to rise, placing considerable pressure on health and social care services (McKee et al., 2021). Physical function, encompassing muscle strength, muscle mass, and functional mobility, is a key determinant of quality of life in older adults (Petnehazy et al., 2024), and impairments in this domain are independently associated with increased mortality (Landi et al., 2016). Declines in physical function are also closely linked to the development of frailty and increased risk of falls (Smee et al., 2012), both of which contribute substantially to hospitalisation and premature mortality in older adults (OHID, 2022). Given this burden, identifying modifiable risk factors that can attenuate age-related declines in physical function and prevent falls across the life course is a public health priority.

Diet is a potentially modifiable factor influencing physical function. At the nutrient level, adequate protein intake has received considerable attention for its role in preserving muscle mass and strength, with higher protein intake positively associated with both fat-free mass and grip strength in a large cross-sectional study of UK Biobank participants (Celis-Morales et al., 2018). Beyond individual nutrients, consumption of specific food groups, such as fruit and vegetables, dairy, and oily fish, have been associated with better functional outcomes including maintained grip strength and preserved muscle mass (Granic et al., 2020; Robinson et al., 2008). More recently, research has shifted towards examining overall dietary patterns, which better capture the complexity of habitual diet and the synergistic effects of foods consumed together. Adherence to patterns such as the Mediterranean diet has been prospectively associated with better physical performance (Silva et al., 2018), and reduced frailty (Kojima et al., 2018) suggesting that overall diet quality may be particularly important for healthy ageing.

In the UK, healthy eating recommendations are visually communicated through the Eatwell Guide, a public-facing tool which promotes consumption of a dietary pattern rich in fruits and vegetables, wholegrains, lean proteins, and dairy, whilst limiting foods high in saturated fat, salt, and sugar (OHID, 2016; Scarborough et al., 2016). Designed to reflect foods familiar and accessible to the UK population, the Eatwell Guide represents the principal dietary framework underpinning UK nutrition policy and practice (Shannon et al., 2024). Whilst evidence linking adherence to health outcomes has only recently begun to emerge, higher Eatwell Guide adherence has been associated with reduced all-cause mortality (Scheelbeek et al., 2020), increased life expectancy (Fadnes et al., 2023), and improvements in markers of adiposity (Griffiths et al., 2025a) and cardiometabolic health (Gregory et al., 2024). However, to date no study has examined associations between Eatwell Guide adherence and markers of physical function, representing a notable gap in the evidence base.

The present study therefore aims to examine both cross-sectional and prospective associations between adherence to the Eatwell Guide and markers of physical function (grip strength, fat-free mass, self-reported walking pace, and falls) within the UK Biobank cohort. These findings will provide valuable insight into the suitability of UK dietary guidelines in supporting healthy ageing and maintaining functional independence in the population.

## Methods

### Study population and design

The UK Biobank is a large prospective cohort study designed to investigate the genetic, lifestyle, and environmental determinants of disease in the UK population, with full details of the study design and methods reported elsewhere (Ollier et al., 2005; Sudlow et al., 2015). Briefly, over 500,000 participants aged 37–73 years were recruited between 2006 and 2010 via NHS patient registers across England, Scotland, and Wales. At the baseline assessment visit, participants completed a touchscreen questionnaire capturing sociodemographic characteristics (e.g., sex, ethnicity, education), lifestyle behaviours including diet (short FFQ questionnaire) and physical activity, and general health. A verbal interview was also conducted, and participants provided biological samples alongside objective measures of physical function. Since baseline, the UK Biobank has been expanded to include repeat dietary assessments (using Oxford WebQ), imaging data, and a range of additional health-related outcomes across multiple follow-up instances.

Ethical approval for the UK Biobank was granted by the North West–Haydock Research Ethics Committee (REC reference: 16/NW/0274), and written electronic informed consent was obtained from all participants.

### Dietary assessment and calculation of Eatwell Guide scores

The present study used dietary intake data that were collected using the Oxford WebQ, a validated web-based 24-hour dietary recall tool developed for use in large-scale epidemiological studies (Greenwood et al., 2019; Liu et al., 2011). The Oxford WebQ captures consumption of 206 food items and 32 beverages over the preceding 24 hours, with participants selecting the number of standard portions consumed for each item. Participants recruited from April 2009 onwards completed the Oxford WebQ at their baseline assessment visit, and those who provided a valid email address were subsequently invited to complete up to four additional recalls administered at approximately three-to-four-month intervals. Participants who completed at least one Oxford WebQ were included. Where multiple dietary assessments were available, values were averaged across recalls to better represent habitual dietary intake. Dietary recalls self-reported as atypical were excluded from all analyses.

### Eatwell Guide adherence score

Adherence to the Eatwell Guide was quantified using a food-based scoring system described in detail elsewhere (Griffiths et al., 2025b). The scoring approach is anchored in foods rather than nutrients, in keeping with the structure of the Eatwell Guide, and is designed to provide actionable public health guidance whilst enabling tracking of dietary changes over time (Willett, 2013).

The scoring tool comprised twelve components: starchy carbohydrates, wholegrains, beans and pulses, fish, white meat, nuts, eggs, fruit and vegetables, dairy, red and processed meat, discretionary foods, and fluid intake (eTable 1). Each component was scored on a scale of 0 to 5 points according to the degree of alignment with Eatwell Guide recommendations, giving a maximum possible total score of 60 points. Full adherence to the recommended intake for a given component was awarded 5 points. For components considered beneficial to health, such as fruit and vegetables, wholegrains, and fish, scores decreased proportionally with lower consumption, with 0 points assigned where intake fell below half the recommended level. For components considered less healthy, including red and processed meat and discretionary foods, 0 points were assigned where consumption exceeded 1.5 times the recommended upper limit. Full rationale for the dietary scoring approach can be found in Griffiths et al. (2025b).

### Outcome assessment

Grip strength and fat-free mass were assessed in line with UK Biobank protocols at baseline and subsequent follow up visits (Sudlow et al., 2015). Grip strength was measured in both the left and right hand using a calibrated, hand dynamometer which measures grip force isometrically. For the present study, and in line with previous research (Celis-Morales et al., 2018), a maximum grip strength across the left and right hand was identified for inclusion in the present analysis. Fat-free mass was derived from bioelectrical impedance analysis (BIA) using the Tanita BC418MA device. BIA-derived fat-free mass has demonstrated strong agreement with DXA-derived measures in UK Biobank participants (Feng et al., 2024), supporting its validity for use in large-scale epidemiological research.

Self-reported walking pace was collected at baseline and follow up visits using the online touchscreen questionnaire. Participants were asked “How would you describe your usual walking pace? Slow; Steady/Average; Brisk; None of the above; Prefer not to answer”. Those participants who answered with ‘none of the above’ or ‘prefer not to answer’ were removed from the analysis. Self-reported walking pace has been shown to be a valid marker of measured walking speed and a useful indicator of physical function in older adults (Syddall et al., 2015). Given the smaller number of participants reporting a slow pace at follow-up, responses were dichotomised into brisk versus non-brisk (slow or steady/average) for analyses, consistent with evidence that brisk walking confers distinct health benefits over slower paces (Yates et al., 2017).

Falls in the last year were also collected at baseline and follow up visits using the online touchscreen questionnaire. Participants were asked “In the last year have you had any falls? No falls; One fall; More than one fall; None of the above; Prefer not to say”. Those who answered ‘none of the above’ or ‘prefer not to say’ were excluded from the analysis. Falls in the last year was operationalised as a binary outcome (one or more falls vs. no falls). This 12-month recall period is supported by evidence demonstrating high specificity (95-96%) and reasonable sensitivity (80-89%) for falls recall in community-dwelling older adults (Ganz et al., 2005). This approach is also suitable given the number of participants at each follow up timepoint, maximising statistical power.

### Statistical analysis

All analyses were conducted in R Studio (version 4.4.0). Overall and stratified (by level of adherence to the Eatwell Guide) baseline characteristics of the analytic sample were summarised as mean ± standard deviation (SD) for continuous variables and as number and percentage of participants for categorical variables. To determine cross-sectional associations between Eatwell Guide adherence and continuous markers of physical function (grip strength, fat-free mass percentage), linear regression analysis was conducted. For categorical markers (self-reported walking pace, self-reported falls in the last year), logistic regression analysis was conducted.

Linear mixed model analyses were conducted to assess prospective associations between Eatwell Guide adherence score and continuous markers of physical function (grip strength, fat-free mass percentage), and Generalised Estimating Equations (GEE) model analysis was conducted for categorical outcomes (self-reported walking pace, self-reported falls in the last year). These analyses were conducted with Eatwell Guide as a continuous score variable, and also split into study-specific tertiles of adherence (low, medium, high). For tertile analyses, the low adherence tertile was used as the reference. For GEE models, the main effect of Eatwell Guide score represents the overall association with the outcome across all timepoints, while the interaction term (Eatwell Guide score × time) tests whether this association changes over follow-up. Analyses were adjusted simultaneously for: age, sex, ethnicity (White, other ethnic background), socio-economic status (Townsend Index categorised as least deprived [quintile 1], middle deprived [quintiles 2–4], most deprived [quintile 5]), education (higher [college/university/other professional qualification], vocational [NVQ/HND/HNC], upper secondary [A-levels], lower secondary [O-levels/GCSEs/CSEs] or none), smoking status (never, previous, current), physical activity (short-form International Physical Activity Questionnaire [IPAQ] group, categorised as low, moderate, high) and energy intake (kcal/day). BMI was adjusted for in models for grip strength, walking pace and falls, but not for fat-free mass percentage as this variable is already expressed relative to total body weight. For visualisation of model-predicted trajectories, predicted values and 95% confidence intervals for each Eatwell Guide tertile (low, medium, high) across all timepoints were derived from the linear mixed-effects models, adjusting for all covariates. To contextualize prospective effect sizes, we expressed differences between Eatwell Guide adherence tertiles as aging equivalents by comparing interaction coefficients (tertile × time) with the age coefficient from cross-sectional models. To contextualize prospective effect sizes, we compared the Eatwell Guide tertile × time interaction coefficients with the age coefficient within the same prospective models. This allowed us to express dietary effects as aging equivalents.

Stratified analyses were conducted to determine whether prospective associations between adherence to the Eatwell Guide and frailty markers were moderated by sex (male vs. female), age (grouped as ‘younger’ vs. ‘older’ by dichotomising at the median (57 years)), physical activity (low vs. moderate vs. high), and protein intake (low vs. medium vs. high tertiles, corresponding to mean intakes of 0.73, 1.05, and 1.46 g/kg/day respectively). Three-way interaction terms (Eatwell Guide score × time × subgroup) were included in linear mixed models (for grip strength and fat-free mass percentage) and generalised estimating equations (for walking pace and falls) to test for effect modification.

### Sensitivity analysis

To assess robustness, sensitivity analyses were conducted: (1) excluding the low-participation first repeat assessment timepoint (Instance 1), (2) restricting to participants with ≥2 dietary recalls, (3) excluding participants with implausible energy intakes (<800 or >4200 kcal/d for males; <600 or >3500 kcal/d for females), (4) leave-one-component-out analyses to identify potential Eatwell Guide components driving associations, and (5) as a validation analysis (for fat-free mass percentage), we examined prospective associations between baseline Eatwell Guide adherence and DXA-derived lean mass percentage at instance 2 (n=19,646) using linear regression, adjusted for the same covariates as the main analysis (excluding BMI).

## Results

### Cohort characteristics

Of the 502,526 participants who underwent baseline assessment in the UK Biobank study, up to 157,457 participants with available data on dietary intake derived from completion of at least one Oxford WebQ and physical function measures were included in prospective analyses. Baseline characteristics of participants with complete baseline data (n=157,348) are provided in Table 1, overall and stratified by Eatwell Guide adherence tertile (low, medium, high).

**Table 1.**
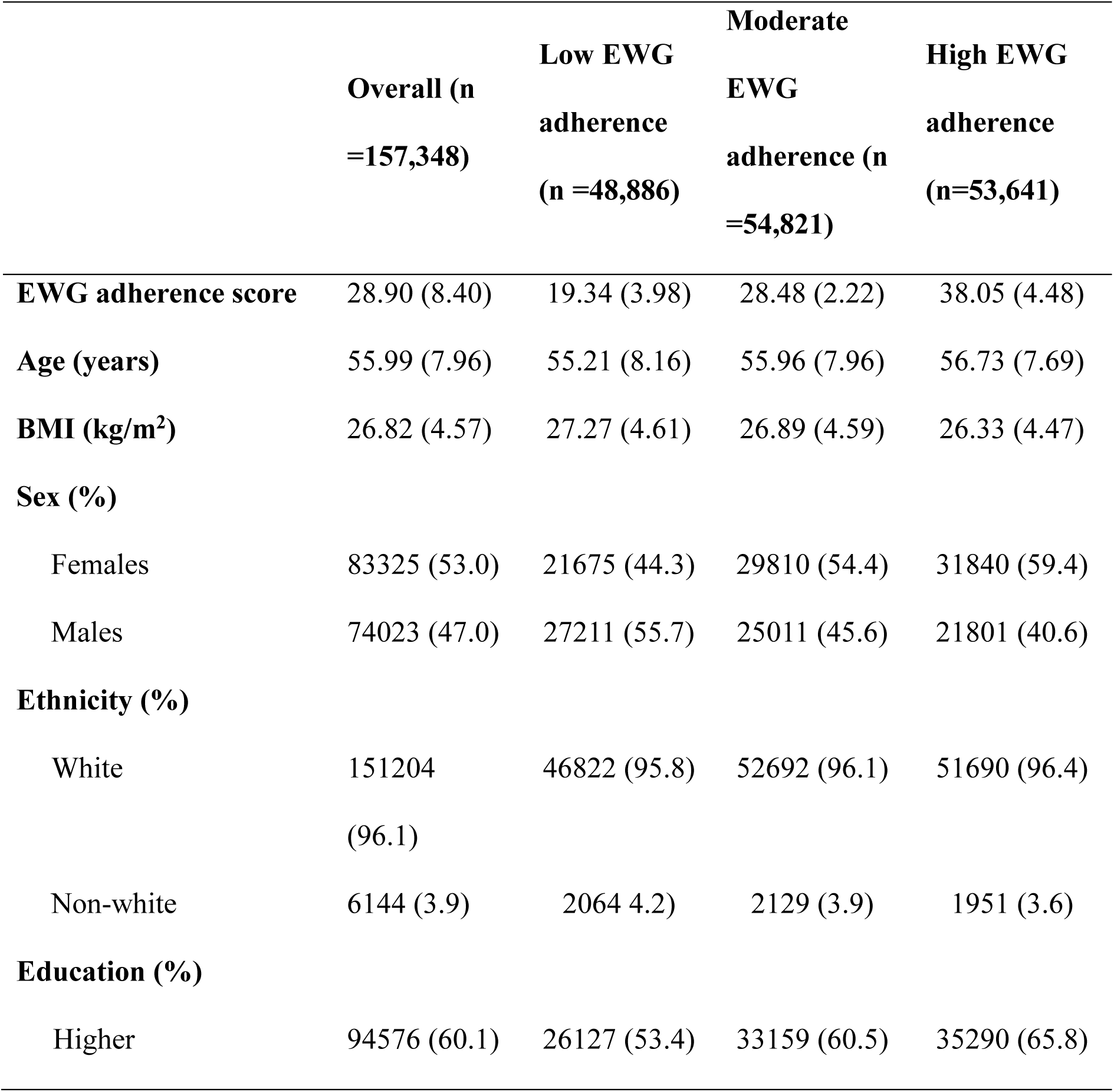
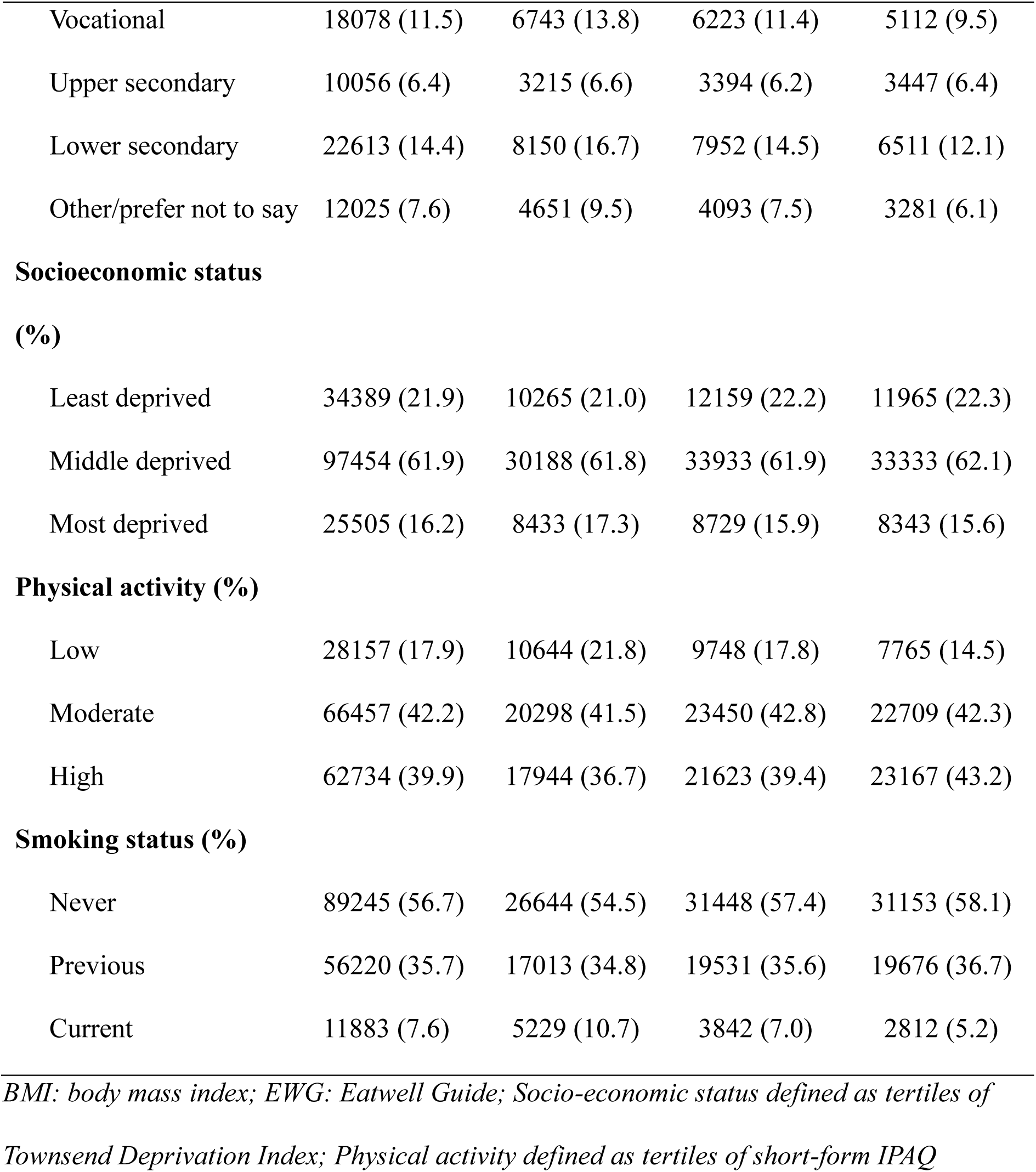
Baseline characteristics of analytical sample.

### Cross-sectional associations between Eatwell Guide adherence and markers of physical function

In cross-sectional analyses, higher adherence to the Eatwell Guide was associated with higher grip strength, higher fat-free mass percentage, higher odds of brisk walking pace, and lower odds of falls in the last year (all p<0.001, eTables 2-3).

In analyses comparing adherence tertiles, moderate and high Eatwell Guide adherence were associated with better physical function markers relative to low adherence (eTables 4-5). Compared with low adherence, high adherence was associated with 0.37 kg higher grip strength, 1% higher fat-free mass percentage, 34% higher odds of brisk walking pace (OR=1.34, 95% CI: 1.30-1.37), and 6% lower odds of experiencing falls in the last year (OR=0.94, 95% CI: 0.91-0.97) (all p<0.001).

### Prospective associations between Eatwell Guide adherence and markers of physical function

In the prospective linear mixed model analysis, higher Eatwell Guide adherence was associated with slower decline in fat-free mass percentage (p<0.001) and attenuation of grip strength decline (p<0.01) (Table 2).

**Table 2.** Linear mixed model analysis of prospective associations between Eatwell Guide adherence score and markers of physical function.

| Predictor (EWG score x time) | B (SE) | P |
| --- | --- | --- |
| Grip strength (kg) (n = 157,457) | 0.005 (0.002) | <0.01 |
| Fat free mass (%) (n = 155,980) | 0.015 (0.001) | <0.001 |

*Models were adjusted for age, sex, ethnicity, socioeconomic status (Townsend Index), education, smoking status, physical activity, and energy intake. Body mass index (BMI) was additionally adjusted for in the grip strength model but not for fat-free mass percentage, as this variable is inherently size-adjusted.*

In the generalised estimating equations analysis of categorical outcomes (Table 3), higher Eatwell Guide adherence was associated with greater odds of brisk walking pace across all timepoints (OR=1.02 per point, 95% CI: 1.02-1.02, p<0.01), though this positive association attenuated over follow-up (EWG*Time interaction OR=0.998, 95% CI: 0.997-0.999, p=0.002). Higher Eatwell Guide adherence was prospectively associated with lower falls risk (OR=0.996 per point, 95% CI: 0.995-0.998, p<0.001), with this protective association remaining stable over time (EWG*Time interaction: p=0.89).

**Table 3.** Generalised estimated equations model analysis of prospective associations between Eatwell guide adherence score and categorical markers of physical function.

| Predictor | OR (Lower CI – Upper CI) | P |
| --- | --- | --- |
| **Self-reported walking pace (n =157,218)** |  |  |
| Main effect (EWG score) | 1.019 (1.017-1.021) | <0.01 |
| Interaction (EWG score * Time) | 0.998 (0.997-0.999) | <0.001 |
| **Self-reported falls in last year (n =157,128)** |  |  |
| Main effect (EWG score) | 0.996 (0.995-0.998) | <0.001 |
| Interaction (EWG score * Time) | 1.000 (0.998-1.000) | 0.89 |

**Note.** Main effect represents the overall association between Eatwell Guide adherence and outcome across all timepoints. Interaction term (EWG score × time) indicates whether this association changes over follow-up. Reference categories: non-brisk walking pace and no falls. Models were adjusted for age, sex, ethnicity, socioeconomic status (Townsend Index), education, BMI, smoking status, physical activity and energy intake.

Analyses comparing adherence tertiles showed clear dose-response relationships for fat-free mass percentage (medium vs low: β=0.123, p<0.001; high vs low: β=0.289, p<0.001), while grip strength showed significant effects only in the highest compared with lowest tertile (β=0.084, p=0.046) (eTable 6). To contextualize effect sizes, adherence in the highest versus lowest tertile was associated with attenuated fat-free mass decline equivalent to approximately 2.5 years less aging, while the grip strength effect was substantially smaller, equivalent to approximately 3.5 months less ageing. Walking pace showed positive main effects for both moderate (OR=1.17, p<0.001) and high (OR=1.35, p<0.001) tertiles, though these advantages attenuated over time (EWG*Time interaction OR=0.972, p=0.032 and OR=0.958, p<0.001, respectively). Falls showed a protective association only in the highest tertile (OR=0.94, p<0.001), with no significant effect in the medium tertile (OR=0.98, p=0.164), and no change in these associations over time (eTable 7). Trajectories of continuous physical function markers over time by Eatwell Guide adherence tertile are shown in Figure 1.

**Figure 1.**
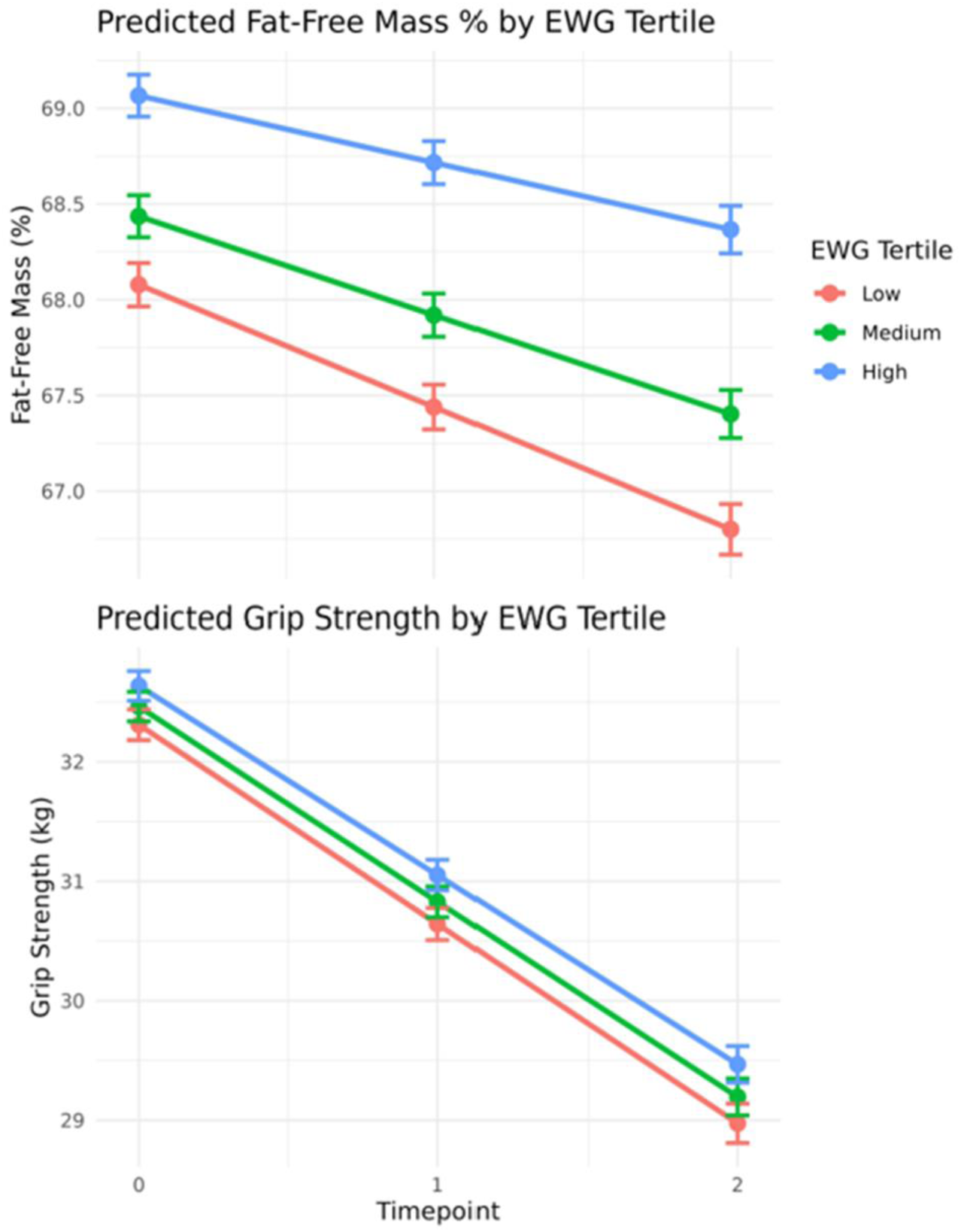
Trajectories of continuous physical function markers over time by Eatwell Guide adherence tertile.

### Sub-group analyses

Interaction tests were conducted to examine whether associations between EWG adherence and prospective outcomes differed by sex, age, or physical activity level. No significant interactions were observed for any outcome by sex (all p>0.4), age (all p>0.17), or physical activity level (all p>0.59), indicating that associations were consistent across these subgroups.

To examine whether protein intake modified associations, we tested three-way interactions (EWG × time × protein tertile). No significant interactions were observed for any outcome (all p>0.1), indicating that EWG associations with muscle mass and function trajectories were consistent across low (0.73 g/kg/day), medium (1.05 g/kg/day), and high (1.46 g/kg/day) protein intake levels.

### Sensitivity analysis

Sensitivity analyses excluding timepoint 1 yielded consistent results, with all significant associations remaining significant (grip strength p=0.002, fat free mass percentage p<0.001, walking pace p<0.001) and falls remaining non-significant (p=0.97). Findings remained robust when restricted to participants with ≥2 dietary recalls (eTable 8) and when excluding extreme energy intake reporters (eTable 9), with all significant associations maintaining significance and direction. Leave-one-component-out analyses revealed that the prospective fat-free mass percentage association was robust regardless of which component was removed (all p<0.001, eTable 10). In contrast, the grip strength association was attenuated when the red meat (p=0.156) and fluid (p=0.07) components were removed (eTable 11). For self-reported walking pace (eTable 12) findings remained significant and for falls (eTable 13) findings remained non-significant with one component removed. In a subset of participants with DXA measurements at instance 2 (n=19,646), higher baseline Eatwell Guide adherence was prospectively associated with greater lean mass percentage (β=0.083%, SE=0.006, p<0.001), providing convergent evidence from gold-standard methodology for the protective association between dietary quality and fat-free mass.

## Discussion

This study represents the first cross-sectional and prospective analysis examining associations between Eatwell Guide adherence and markers of physical function. Cross-sectionally, higher Eatwell Guide adherence was associated with higher muscle mass, grip strength, greater odds of brisk walking pace, and lower odds of falls. Prospectively, higher Eatwell Guide adherence was associated with attenuated muscle mass decline, with a clear dose-response relationship observed across adherence tertiles. Grip strength showed significant but weaker attenuations over time, with effects observed only in the highest, compared with the lowest adherence tertile. For functional mobility outcomes, higher adherence was prospectively associated with substantially higher odds of brisk walking pace (35% higher odds in highest vs lowest tertile) and modestly lower falls risk (6% lower odds). However, while the falls association remained stable over time, the walking pace advantage attenuated during follow-up. These findings suggest that adherence to the Eatwell Guide confers clear muscle mass-protective benefits, alongside modest but consistent effects on functional strength and falls risk. However, the attenuation of walking pace advantages over time demonstrates that dietary quality alone cannot prevent age-related functional decline.

The most consistent and robust finding in the present study was the prospective association between higher Eatwell Guide adherence and attenuated fat-free mass decline, which was equivalent to approximately 2.5 years less ageing in the highest compared to the lowest adherence tertile. The DXA validation subsample corroborated this finding, with higher baseline Eatwell Guide adherence associated with greater lean mass percentage at follow-up. Prospective evidence linking adherence to other dietary patterns, such as the Mediterranean diet, to muscle mass trajectories remains limited but typically reports positive associations (Dominguez et al., 2025). The present findings extend this evidence base by demonstrating a prospective association between a UK-specific dietary framework and attenuated fat-free mass decline in a large general population cohort. The Eatwell Guide promotes several food groups with established myoprotective properties, including diverse protein sources, fruit and vegetables, dairy, and fish, which may collectively support muscle protein synthesis and attenuate inflammation-driven catabolism through antioxidative and anti-inflammatory pathways (Granic et al., 2020). The absence of any significant interaction by protein intake tertile indicates that the score’s protective association is not confined to those with higher protein intakes and may reflect the broader dietary quality captured by the index.

Both cross-sectional and prospective associations between Eatwell Guide adherence and grip strength were statistically significant, though modest in magnitude. Cross-sectionally, the finding is consistent with evidence linking dietary quality and/or specific food groups (e.g., oily fish, fruit and vegetables, and red meat) to muscle strength in older adults (Gedmantaite et al., 2020; Robinson et al., 2008). The prospective effect was equivalent to approximately 3.5 months less ageing in the higher compared with lower adherence groups, a modest magnitude that may reflect the broader range of determinants of grip strength beyond nutritional intake, including neuromuscular coordination, physical activity, and acute health status (Carson, 2018; McGregor et al., 2014). These findings align with some prospective evidence on the Mediterranean diet, which found no significant association with grip strength (Huang et al., 2021; Talegawkar et al., 2012), suggesting that adherence to the Eatwell Guide may capture a small but detectable dietary contribution to grip strength that has not been consistently demonstrated for other dietary patterns.

Cross-sectionally, higher Eatwell Guide adherence was associated with 34% greater odds of brisk walking in the highest versus lowest adherence tertile at baseline. However, the prospective interaction effects indicated that the baseline advantage diminishes over time, suggesting that adherence to the Eatwell Guide may be insufficient alone to prevent the age-related decline in walking pace. This likely reflects the multifactorial nature of walking pace in older adults, which is shaped by the complex interaction of muscular strength, balance, proprioception, cardiovascular fitness, and physical activity - factors that dietary quality alone may be insufficient to modify (Hainline et al., 2024). These findings extend prospective evidence from a British birth cohort, in which higher diet quality was associated with faster walking speed seven years later in women but not men (Tektonidis et al., 2020). While we similarly observed positive associations between Eatwell Guide adherence and brisk walking pace, our repeated-measures design revealed that this advantage attenuates over time, and that dietary quality does not prevent the age-related decline observed in both males and females. Our findings suggest that diet quality it is insufficient as a standalone intervention to prevent mobility decline in aging populations.

Higher Eatwell Guide adherence was prospectively associated with lower falls risk, and this protective association remained stable over time. Tertile analyses revealed that the protective association was significant only in the highest vs. lowest adherence group suggesting a potential threshold effect whereby benefits may require relatively high adherence to the Eatwell Guide. While statistically significant, the modest magnitude of this effect (6% lower odds in the highest vs lowest tertile) indicates a small protective benefit. Given that dietary effects on falls risk are likely to operate indirectly through maintenance of muscle mass and strength, the modest prospective associations observed for these outcomes may explain these findings. This is in contrast to prospective evidence from the Mediterranean diet literature, where higher adherence was associated with approximately 28% lower odds of falling in community-dwelling older Spanish adults (Ballesteros et al., 2020). The Mediterranean diet’s emphasis on anti-inflammatory foods such as olive oil, may partly explain this discrepancy, given the proposed role of chronic inflammation in sarcopenia and falls risk (Ballesteros et al., 2020). However, this is unlikely to fully explain the large differences in the magnitude of findings.

This study has several notable strengths. The large sample size provided substantial statistical power to detect associations between dietary quality and physical function outcomes, and to examine interaction effects over time. The prospective design with repeated outcome assessment allowed examination of both cross-sectional associations and longitudinal change, enabling a more nuanced interpretation of the direction and temporal nature of observed effects and reducing risk of reverse causality. The Eatwell Guide score used to assess diet quality is directly relevant to public health policy in this country and represents an advance over prior work in this area that has largely relied on Mediterranean diet and general healthy diet indices with limited applicability to UK dietary patterns. In addition, the analyses produced broadly consistent effects when individual components were removed, suggesting that associations are unlikely to be driven by one individual component but instead suggest that they are due to overall diet quality. Several limitations should be acknowledged. The observational design limits causal inference. Self-reported dietary intake is subject to measurement error and social desirability bias, though previous work has demonstrated reasonable stability of dietary reporting in UK Biobank (Bradbury et al., 2018). Self-reported falls and walking pace introduce potential recall bias and misclassification, which may have reduced our ability to detect associations. The UK Biobank cohort exhibits healthy volunteer bias, with participants on average more affluent, well-educated and less ethnically diverse than the general UK population, limiting generalisability (Fry et al., 2017). Residual confounding can also not be excluded, as physical activity may function as a mediator rather than simply a confounder, particularly for walking pace, potentially attenuating prospective associations. The binary coding of falls and walking pace may have reduced sensitivity to detect associations by not capturing the full gradient of variation, although this classification was considered necessary to ensure sufficient sample size in each analytical group. Also, the translation of qualitative Eatwell Guide recommendations into quantitative scoring criteria required interpretative decisions, and alternative approaches may yield different results.

The findings of this study have implications for public health nutrition policy in the UK. The prospective associations observed between Eatwell Guide adherence and attenuated decline in fat-free mass, alongside a modest effect on grip strength, demonstrate that alignment with current UK dietary recommendations may confer some benefit for musculoskeletal health in older adults. The absence of significant effect modification by age, sex, or BMI suggests that these benefits may be broadly consistent across population subgroups. However, our inability to conduct subgroup analyses by socioeconomic status or ethnicity, due to the predominantly White (96%) and relatively affluent sample, represents an important limitation. Previous qualitative research has highlighted substantial variation in dietary beliefs, knowledge, and engagement with the Eatwell Guide across different socioeconomic and ethnic groups (Maguire and Monsivais, 2015; Ojo et al., 2023) raising questions about the generalisability of our findings to more diverse and deprived populations. While modest protective effects for falls risk were observed, the attenuation of the walking pace advantage over time demonstrates that dietary quality alone is unlikely to be sufficient to prevent broader functional decline, and that Eatwell Guide adherence is best considered one component of a multifaceted approach to healthy ageing alongside physical activity promotion and other lifestyle behaviours.

In conclusion, this large-scale prospective cohort study demonstrated that adherence to the Eatwell Guide was associated with attenuated decline in fat-free mass, modest preservation of grip strength, and lower falls risk, with no significant effect modification across key demographic subgroups. While higher adherence was also associated with better walking pace, this advantage attenuated over follow-up, indicating that dietary quality alone does not prevent age-related decline in walking speed. These findings suggest that alignment with current UK dietary recommendations may support musculoskeletal health and reduce falls risk in older adults, though the modest effect sizes for grip strength and falls, as well as the attenuation of walking pace advantages highlight that diet quality should be considered one component of a multifaceted approach to preventing functional decline in aging populations. Further research is needed to explore the mechanisms underlying these associations and to evaluate whether interventions to improve Eatwell Guide adherence in older adults can translate to meaningful gains in physical function and independence. It should also be determined whether these associations replicate in more diverse populations, including evaluation of culturally adapted dietary guidelines such as the African-Caribbean (Saint Hill, 2023) and South Asian (Jay, 2021) Eatwell Guides, to ensure that dietary recommendations are equitable and relevant across all UK population groups.

## Data Availability

Data used for this study were obtained from the UK Biobank and are subject to access restrictions.

## Acknowledgements

This research has been conducted using the UK Biobank Resource under application number 147784.

## Availability of Data and Materials

Data used for this study were obtained from the UK Biobank and are subject to access restrictions.

