## Supplementary materials for "Adherence to the Eatwell Guide and associations with markers of physical function: A prospective analysis within the UK Biobank cohort"

### List of supplementary tables

**Supplementary eTable 1.** List of individual foods which contributed to each food group and the Eatwell Guide scoring methodology

**Supplementary eTable 2.** Linear regression of associations between adherence to the Eatwell Guide and markers of physical function

**Supplementary eTable 3.** Logistic regression analysis of associations between adherence to the Eatwell Guide and markers of physical function

**Supplementary eTable 4**. Linear regression analysis of associations between Eatwell Guide adherence tertiles and continuous markers of physical function

**Supplementary eTable 5**. Logistic regression of associations between EWG adherence tertiles and categorical markers of physical function

**Supplementary eTable 6**. Linear mixed model analysis of prospective associations between Eatwell Guide adherence tertiles and markers of physical function

**Supplementary eTable 7.** Generalised estimated equations model analysis of prospective associations between Eatwell guide adherence tertiles and categorical markers of physical function

**Supplementary eTable 8**. Sensitivity analysis of prospective associations between Eatwell Guide adherence and markers of physical function, but limited to participants with a minimum of two dietary reports

**Supplementary eTable 9**. Sensitivity analysis of prospective associations between Eatwell Guide adherence and markers of physical function, but excluding participants with extreme energy intakes

**Supplementary eTable 10**. Sensitivity analysis of prospective associations between Eatwell Guide adherence and fat free mass percentage, but with one component removed

**Supplementary eTable 11**. Sensitivity analysis of prospective associations between Eatwell Guide adherence and grip strength, but with one component removed

**Supplementary eTable 12**. Sensitivity analysis of prospective associations between Eatwell Guide adherence and self-reported walking pace, but with one component removed

**Supplementary eTable 13.** Sensitivity analysis of prospective associations between Eatwell Guide adherence and falls, but with one component removed

**Supplementary eTable 1.** List of individual foods which contributed to each food group and the Eatwell Guide scoring methodolog

| Food component | Contributing foods from the Oxford WebQ | Average consumption required. | EWG Score |
|---|---|---|---|
| **Fruit and vegetables** | Stewed fruit (104410), Prune (104420), Dried fruit (104430), Mixed fruit (104440), Apple (104450), Banana (104460), Berry (104470), Cherry (104480), Grapefruit (104490), Grape (104500), Mango (104510), Melon (104520), Orange (104530), Satsuma (104540), Peach nectarine (104550), Pear (104560), Pineapple (104570), Plum (104580), Other fruit (104590), Orange juice (100190), Grapefruit juice (100200), Pure fruit vegetable juice (100210), Fruit smoothie (100220), Mixed veg (104060), Veg pieces (104070), Coleslaw (104080), Side salad (104090), Avocado (104100), Beetroot (104130), Broccoli (104140), Butternut squash (104150), Cabbage kale (104160), Carrot (104170), Cauliflower (104180), Celery (104190), Courgette (104200), Cucumber (104210), Garlic (104220), Leek (104230), Lettuce (104240), Mushroom (104250), Onion (104260), Parsnip (104270), Sweet pepper (104290), Spinach (104300), Sprouts (104310), Sweetcorn (104320), Fresh tomato (104340), Tinned tomato (104350), Turnip swede (104360), Watercress (104370), Other veg (104380), Olive (102490) | From 0 to < 2.5 | 0 |
| | | From 2.5 to < 3.125 | 1 |
| | | From 3.125 to < 3.75 | 2 |
| | | From 3.75 to < 4.375 | 3 |
| | | From 4.375 to < 5 | 4 |
| | | ≥5 | 5 |
| **Starchy Carbohydrates** | White pasta (102710), Wholemeal pasta (102720), White rice (102730), Brown rice (102740), Snackpot (102760), Couscous (102770), Other grain (102780), Sliced bread (100950), Baguette (101020), Bap (101090), Bread roll (101160), Naan bread (101230), Garlic bread (101240), Crispbread (101250), Oatcake (101260), Other bread (101270), Porridge (100770), Muesli (100800), Oat crunch (100810), Plain cereal (100830), Bran cereal (100840), Wholewheat cereal (100850), Other cereal (100860), Fried potatoes (104020), Boiled baked potatoes (104030), Mashed potato (104050), Sweet potato (104330) | **Men** < 2.5 **Women** < 2 | 0 |
| | | **Men** From 2.5 to < 3.125 **Women** From 2 to < 2.5 | 1 |
| | | **Men** From 3.125 to < 3.75 | 2 |

| | | | |
|---|---|---|---|
| | | **Women**<br>From 2.5 to < 3 | |
| | | **Men**<br>From 3.75 to < 4.375<br>**Women**<br>From 3 to < 3.5 | 3 |
| | | **Men**<br>From 4.375 to < 5<br>**Women**<br>From 3.5 to < 4 | 4 |
| | | **Men**<br>≥ 5<br>**Women**<br>≥ 4 | 5 |
| **Wholegrains** | Wholemeal pasta (102720), Brown rice (102740), Sliced bread (100950), Type of bread (20091), Baguette (101020), Type of baguette (20092), Bap (101090), Type of bap (20093), Bread roll (101160), Type of bread roll (20094), Oatcake (101260), Porridge (100770), Muesli (100800), Bran cereal (100840), Wholewheat cereal (100850), Other grain (102780) | From 0 to < 1.5 | 0 |
| | | From 1.5 to < 1.875 | 1 |
| | | From 1.875 to < 2.25 | 2 |
| | | From 2.25 to < 2.625 | 3 |
| | | From 2.625 to < 3 | 4 |
| | | ≥ 3 | 5 |
| **Beans/pulses** | | From 0 to < 0.21 | 0 |

| | | | |
|---|---|---|---|
| | Pea (104280), Green beans (104120), Broad beans (104110), Baked beans (104000), Pulses (104010), Tofu (103270) | From 0.21 to < 0.27 | 1 |
| | | From 0.27 to < 0.32 | 2 |
| | | From 0.32 to < 0.38 | 3 |
| | | From 0.38 to < 0.43 | 4 |
| | | ≥ 0.43 | 5 |
| **Fish**[a] | Tinned tuna (103150), Oily fish (103160), Breaded fish (103170), Battered fish (103180), White fish (103190), Prawns (103200), Lobster crab (103210), Shellfish (103220), Other fish (103230) | **Fish**<br>From 0 to < 0.14<br>**Oily fish**<br>From 0 to < 0.07 | 0 |
| | | **Fish**<br>From 0.14 to < 0.18<br>**Oily fish**<br>From 0.07 to < 0.09 | 1 |
| | | **Fish**<br>From 0.18 to < 0.21<br>**Oily fish**<br>From 0.09 to < 0.11 | 2 |
| | | **Fish**<br>From 0.21 to < 0.25<br>**Oily fish**<br>From 0.11 to < 0.13 | 3 |

| | | | |
|---|---|---|---|
| | | **Fish**<br>From 0.25 to < 0.29<br>**Oily fish**<br>From 0.13 to < 0.14 | 4 |
| | | **Fish**<br>≥ 0.29<br>**Oily fish**<br>≥ 0.14 | 5 |
| **Poultry** | Poultry (103060), Breaded poultry (103050) | From 0 to < 0.07 | 0 |
| | | From 0.07 to < 0.09 | 1 |
| | | From 0.09 to < 0.11 | 2 |
| | | From 0.11 to < 0.13 | 3 |
| | | From 0.13 to < 0.14 | 4 |
| | | ≥ 0.14 | 5 |
| **Nuts** | Unsalted nuts (102440), salted nuts (102430), unsalted peanuts (102420), salted peanuts (102410) | From 0 to < 0.07 | 0 |
| | | From 0.07 to < 0.09 | 1 |
| | | From 0.09 to < 0.11 | 2 |
| | | From 0.11 to < 0.13 | 3 |
| | | From 0.13 to < 0.14 | 4 |
| | | ≥ 0.14 | 5 |
| **Eggs** | | From 0 to < 0.07 | 0 |

| | | | |
|---|---|---|---|
| | Whole egg (102940), Omelette (102950), Egg sandwiches (102960), Scotch egg (102970), Other egg (102980) | From 0.07 to < 0.09 | 1 |
| | | From 0.09 to < 0.11 | 2 |
| | | From 0.11 to < 0.13 | 3 |
| | | From 0.13 to < 0.14 | 4 |
| | | ≥ 0.14 | 5 |
| **Red and processed meat** | Bacon (103070), Ham (103080), Liver (103090), Sausage (103010), Beef (103020), Pork (103030), Lamb (103040) | ≥ 1.5 | 0 |
| | | From 1.375 to < 1.5 | 1 |
| | | From 1.25 to < 1.375 | 2 |
| | | From 1.12 To < 1.25 | 3 |
| | | From 1 to < 1.12 | 4 |
| | | < 1 | 5 |
| **Dairy** | Yogurt (102090), Low fat hard cheese (102810), Hard cheese (102820), Low fat cheese spread (102850), Cheese spread (102860), Soft cheese (102830), Goat cheese (102900), Blue cheese (102840), Feta (102880), Mozzarella (102890), Other cheese (102910), Cottage cheese (102870), Milk (100520), Flavoured milk (100530), Added milk instant coffee (100260), Added milk filtered coffee (100280), Added milk espresso (100320), Added milk other coffee (100350), Added milk standard tea (100460), Added milk rooibos tea (100480), Instant coffee (100250), Filtered coffee (100270), Espresso (100310), Other coffee (100330), Standard tea (100400), Rooibos tea (100410), Dairy smoothie (100230), Latte (100300), Cappuccino (100290), Type of milk used (100920), Added milk cereal (100890), Porridge (100770), Muesli (100800), Bran cereal (100840), Wholewheat cereal (100850), Oat crunch (100810), Plain cereal (100830), Sweetened cereal (100820), Other cereal (100860) | ≥ 3 | 0 |
| | | From 2.75to < 3 | 1 |
| | | From 2.5 to < 2.75 | 2 |
| | | From 2.25 to < 2.5 | 3 |
| | | From 2 to < 2.25 | 4 |
| | | < 2 | 5 |
| **Discretionary foods** | Chocolate biscuit (102350), Chocolate covered biscuit (102340), Chocolate bar (102260), Chocolate sweets (102310), Chocolate raisins (102300), Dark chocolate (102290), Milk chocolate (102280), White chocolate (102270), Sweet biscuits (102360), Cakes (102190), Cheesecake (102220), Doughnut (102200), Fruitcake (102180), Danish pastry (102060), Sponge pudding (102210), Milk based pudding (102140), Other milk pudding (102150), | ≥ 1.5 | 0 |
| | | From 1.375 to < 1.5 | 1 |
| | | From 1.25 to < 1.375 | 2 |

| | | | |
|---|---|---|---|
| | Other desert (102230), Soya desert (102170), Sweets (102330), Diet sweets (102320), Other sweets (102380), Ice cream (102120), Fizzy drinks (100170), Squash (100180), Sugar added to tea (100490), Sugar added to coffee (100370), Sugar added to cereal (100900), Hot chocolate (100550), Pancake (102010), Scotch pancake (102020), Croissant (102050), Scone (102070), Crisp (102460), Cereal bar (102370) | From 1.12 to < 1.25 | 3 |
| | | From 1 to < 1.12 | 4 |
| | | < 1 | 5 |
| **Fluid** | Drinking water (100150), Instant coffee (100250), Filtered coffee (100270), Espresso (100310), Cappuccino (100290), Latte (100300), Other coffee type (100330), Decaffeinated coffee (100360), Standard tea (100400), Rooibos tea (100410), Green tea (100420), Herbal tea (100430), Other tea (100440), Decaffeinated tea (100470), Low calorie drink (100160), Squash (100180), Orange juice (100190), Grapefruit juice (100200), Pure fruit vegetable juice (100210), Fruit smoothie (100220), Dairy smoothie (100230), Type of milk used (100920), Milk (100520), Flavoured milk (100530) | From 0 to < 3 | 0 |
| | | From 3 to < 3.75 | 1 |
| | | From 3.75 to < 4.5 | 2 |
| | | From 4.5 to < 5.25 | 3 |
| | | From 5.25 to < 6 | 4 |
| | | ≥ 6 | 5 |

**Supplementary eTable 2.** Linear regression of associations between adherence to the Eatwell Guide and markers of physical function

| **Outcome** | **B (SE)** | **P** |
|---|---|---|
| Grip strength (kg) (n = 157,348) | 0.020 (0.002) | <0.001 |
| Fat free mass (%) (n = 155,577) | 0.054 (0.002) | <0.001 |

**Supplementary eTable 3.** Logistic regression analysis of associations between adherence to the Eatwell Guide and markers of physical function

| **Outcome** | **OR (Lower CI – Upper CI)** | **P** |
|---|---|---|
| Self-reported walking pace (n = 157,135) | | |
| Non-brisk (reference) (n = 83,751) | - | - |
| Brisk (n = 73,384) | 1.02 (1.02-1.02) | <0.001 |
| Self-reported falls in last year (n = 157,425) | | |
| No falls (reference) (n = 129,463) | - | - |
| One or more falls (n = 27,962) | 0.996 (0.995-0.998) | <0.001 |

**Supplementary eTable 4**. Linear regression analysis of associations between Eatwell Guide adherence tertiles and continuous markers of physical function

| **Outcome** | **Low (reference)** | | **Medium** | | **High** | |
|---|---|---|---|---|---|---|
| | B (SE) | P | B (SE) | P | B (SE) | P |
| Grip strength (n = 157,348) | - | - | 0.182 (0.045) | <0.001 | 0.373 (0.046) | <0.001 |
| Fat free mass (%) (n = 155,577) | - | - | 0.368 (0.039) | <0.001 | 1.00 (0.040) | <0.001 |

**Supplementary eTable 5**. Logistic regression of associations between EWG adherence tertiles and categorical markers of physical function

| **Outcome** | **Low (reference)** | | **Medium** | | **High** | |
|---|---|---|---|---|---|---|
| | OR (95% CI) | P | OR (95% CI) | P | OR (95% CI) | P |
| Self-reported walking pace (n = 157, 135) | | | | | | |
| Non-brisk (reference) (n = 83,751) | - | - | - | - | - | - |
| Brisk (n = 73,384) | - | - | 1.16 (1.13-1.19) | <0.001 | .1.34 (1.30-1.37) | <.0.001 |
| Self-reported falls in last year (n = 157,425) | | | | | | |
| No falls (reference) (n = 129,463) | - | - | - | - | - | - |
| One or more falls (n = 27,962) | - | - | 0.98 (0.94-1.01) | 0.13 | 0.94 (0.91-0.97) | <0.001 |

**Supplementary eTable 6**. Linear mixed model analysis of prospective associations between Eatwell Guide adherence tertiles and markers of physical function

| **Predictor (EWG score x time)** | **Low (reference)** | | **Medium** | | **High** | |
|---|---|---|---|---|---|---|
| | B (SE) | P | B (SE) | P | B (SE) | P |
| Grip strength (n =157,457) | - | - | 0.034 (0.043) | 0.419 | 0.084 (0.042) | 0.046 |
| Fat free mass (%) (n = 155,980) | - | - | 0.123 (0.026) | <0.001 | 0.289 (0.026) | <0.001 |

**Supplementary eTable 7.** Generalised estimated equations model analysis of prospective associations between Eatwell guide adherence tertiles and categorical markers of physical function

| **Predictor (EWG score x time)** | **Low (reference)** | | **Medium** | | **High** | |
|---|---|---|---|---|---|---|
| | OR (95% CI) | P | OR (95% CI) | P | OR (95% CI) | P |
| Self-reported walking pace (n = 157,218) | | | | | | |
| Main effect (EWG score) | - | - | 1.17 (1.14-1.19) | <0.001 | 1.35 (1.31-1.38) | <0.001 |
| Interaction (EWG score * Time) | - | - | 0.972 (0.984-0.998) | 0.03 | 0.958 (0.933-0.982) | <0.001 |
| Self-reported falls in last year (n = 157,128) | | | | | | |
| Main effect (EWG score) | - | - | 0.98 (0.95-1.01) | 0.16 | 0.94 (0.91-0.97) | <0.001 |
| Interaction (EWG score * Time) | - | - | 1.00 (0.966-1.040) | 0.917 | 0.991 (0.956-1.030) | 0.625 |

**Supplementary eTable 8**. Sensitivity analysis of prospective associations between Eatwell Guide adherence and markers of physical function, but limited to participants with a minimum of two dietary reports

| **Predictor (EWG score x time)** | **B (SE)** | **P** |
|---|---|---|
| Grip strength (kg) (n = 102,292) | 0.007 (0.002) | <0.01 |
| Fat free mass (%) (n = 101,388) | 0.020 (0.001) | <0.001 |
| **Predictor (EWG score x time)** | **OR (Lower CI – Upper CI)** | **P** |
| Self-reported walking pace (n =102,101) | | |
| Non-brisk (reference) | - | - |
| Brisk | 0.998 (0.997-1.000) | 0.01 |
| Self-reported falls in last year (n = 102,164) | | |
| No falls (reference) | - | - |
| One or more falls | 1.000 (0.998-1.000) | 0.99 |

**Supplementary eTable 9**. Sensitivity analysis of prospective associations between Eatwell Guide adherence and markers of physical function, but excluding participants with extreme energy intakes

| **Predictor (EWG score x time)** | **B (SE)** | **P** |
|---|---|---|
| Grip strength (kg) (n = 155,971) | 0.005 (0.002) | <0.001 |
| Fat free mass (%) (n = 154,511) | 0.016 (0.001) | <0.001 |
| **Predictor (EWG score x time)** | **OR (Lower CI – Upper CI)** | **P** |
| Self-reported walking pace (n = 155,741) | | |
| Non-brisk (reference) | - | - |
| Brisk | 0.998 (0.997-0.999) | <0.001 |
| Self-reported falls in last year (n =155,653) | | |
| No falls (reference) | - | - |
| One or more falls | 1.000 (0.998-1.000) | 0.91 |

**Supplementary eTable 10**. Sensitivity analysis of prospective associations between Eatwell Guide adherence and fat free mass percentage, but with one component removed

| Predictor (EWG score x time) | B (SE) | P |
|---|---|---|
| EWG score | | |
| Minus starchy carbohydrate | 0.016 (0.001) | <0.001 |
| Minus wholegrains | 0.016 (0.001) | <0.001 |
| Minus red meat | 0.016 (0.001) | <0.001 |
| Minus fish | 0.016 (0.001) | <0.001 |
| Minus white meat | 0.018 (0.001) | <0.001 |
| Minus fruit and vegetables | 0.014 (0.001) | <0.001 |
| Minus dairy | 0.016 (0.001) | <0.001 |
| Minus beans and pulses | 0.017 (0.001) | <0.001 |
| Minus nuts | 0.016 (0.001) | <0.001 |
| Minus eggs | 0.016 (0.001) | <0.001 |
| Minus discretionary food | 0.014 (0.001) | <0.001 |
| Minus fluid | 0.014 (0.001) | <0.001 |

**Supplementary eTable 11**. Sensitivity analysis of prospective associations between Eatwell Guide adherence and grip strength, but with one component removed

| Predictor (EWG score x time) | B (SE) | P |
|---|---|---|
| EWG score | | |
| Minus starchy carbohydrate | 0.007 (0.002) | <0.01 |
| Minus wholegrains | 0.009 (0.002) | <0.01 |
| Minus red meat | 0.003 (0.002) | 0.16 |
| Minus fish | 0.006 (0.002) | <0.01 |
| Minus white meat | 0.005 (0.002) | 0.03 |
| Minus fruit and vegetables | 0.005 (0.002) | 0.02 |
| Minus dairy | 0.005 (0.002) | <0.01 |
| Minus beans and pulses | 0.006 (0.002) | <0.01 |
| Minus nuts | 0.005 (0.002) | 0.02 |
| Minus eggs | 0.006 (0.002) | <0.01 |
| Minus discretionary food | 0.005 (0.002) | 0.02 |
| Minus fluid | 0.004 (0.002) | 0.07 |

**Supplementary eTable 12**. Sensitivity analysis of prospective associations between Eatwell Guide adherence and self-reported walking pace, but with one component removed

| Predictor (EWG score x time) | OR (lower CI – upper CI) | P |
|---|---|---|
| EWG score | | |
| Minus starchy carbohydrate | 0.998 (0.997-0.999) | <0.001 |
| Minus wholegrains | 0.998 (0.997-0.999) | <0.01 |
| Minus red meat | 0.998 (0.996-0.999) | <0.001 |
| Minus fish | 0.998 (0.997-0.999) | <0.01 |
| Minus white meat | 0.997 (0.996-0.999) | <0.001 |
| Minus fruit and vegetables | 0.998 (0.997-0.999) | <0.01 |
| Minus dairy | 0.997 (0.996-0.999) | <0.001 |
| Minus beans and pulses | 0.998 (0.997-0.999) | <0.01 |
| Minus nuts | 0.998 (0.997-0.999) | <0.01 |
| Minus eggs | 0.998 (0.996-0.999) | <0.001 |
| Minus discretionary food | 0.997 (0.996-0.999) | <0.001 |
| Minus fluid | 0.998 (0.996-0.999) | <0.001 |

**Supplementary eTable 13.** Sensitivity analysis of prospective associations between Eatwell Guide adherence and falls, but with one component removed

| **Predictor (EWG score x time)** | **OR (lower CI – upper CI)** | **P** |
|---|---|---|
| EWG score | | |
| Minus starchy carbohydrate | 1.000 (0.998-1.002) | 0.89 |
| Minus wholegrains | 1.000 (0.998-1.002) | 0.91 |
| Minus red meat | 1.000 (0.998-1.002) | 0.85 |
| Minus fish | 1.000 (0.998-1.002) | 0.96 |
| Minus white meat | 1.001 (0.999-1.002) | 0.56 |
| Minus fruit and vegetables | 1.000 (0.998-1.002) | 0.84 |
| Minus dairy | 1.000 (0.998-1.002) | 0.87 |
| Minus beans and pulses | 1.000 (0.998-1.002) | 0.93 |
| Minus nuts | 1.000 (0.998-1.002) | 0.66 |
| Minus eggs | 1.000 (0.998-1.002) | 0.82 |
| Minus discretionary food | 1.000 (0.998-1.002) | 0.65 |
| Minus fluid | 1.000 (0.998-1.002) | 0.98 |
